# Detection of Colorectal Adenocarcinoma and Grading Dysplasia on Histopathologic Slides Using Deep Learning

**DOI:** 10.1101/2022.09.19.22280112

**Authors:** June Kim, Naofumi Tomita, Arief A. Suriawinata, Saeed Hassanpour

## Abstract

Colorectal cancer is one of the most common types of cancer among men and women. The grading of dysplasia and the detection of adenocarcinoma are important clinical tasks in the diagnosis of colorectal cancer and shape the patients’ follow-up plans. This study evaluates the feasibility of deep learning models for the classification of colorectal lesions into four classes: benign, low-grade dysplasia, high-grade dysplasia, and adenocarcinoma. To this end, we develop a deep neural network on a training set of 655 whole-slide images of digitized colorectal resection slides from a tertiary medical institution and evaluate it on an internal test set of 234 slides, as well as on an external test set of 606 adenocarcinoma slides from The Cancer Genome Atlas database. Our model achieves an overall accuracy, sensitivity, and specificity of 95.5%, 91.0%, and 97.1% on the internal test set and an accuracy and sensitivity of 98.5% for adenocarcinoma detection task on the external test set. Our results suggest that such deep learning models can potentially assist pathologists in grading colorectal dysplasia, detecting adenocarcinoma, prescreening, and prioritizing the reviewing of suspicious cases to improve the turnaround time for patients with a high risk of colorectal cancer. Furthermore, the high sensitivity on the external test set suggests our model’s generalizability in detecting colorectal adenocarcinoma on whole slide images across different institutions.

## Introduction

Colorectal cancer (CRC) is the third most common cancer type among men and the second most common cancer type among women.^1^ CRC usually starts as a polyp in the innermost layer of the colon or rectum and spreads outward. Colorectal polyps can progress to cancer over the course of 10-15 years. However, once CRC develops, it can quickly spread and become metastatic: a 2018 study of Swedish patients found that 93% of cases were diagnosed with liver metastases within 3 years of a CRC diagnosis.^2^ Furthermore, the 5-year survival rate for CRC cancer decreases dramatically with its stage; according to the SEER database, the 5-year survival rate for CRC at the “localized” stage is 91%, while at the “distant” stage, it drops to 14%.^3^ Therefore, timely diagnosis of CRC and its early precursors can be life-saving. The likelihood of future occurrences, or a current presence of colorectal cancer can often be assessed via the degree of *dysplasia*, which are abnormal cells that have not yet developed into cancer. One study found that the risk of polyps with high-grade dysplasia (HGD) harboring cancer was 35%, compared to 6% for those with low-grade dysplasia (LGD).^4^

Therefore, the determination of the existence of adenocarcinoma, if none are present, or the varying degrees of dysplasia, is an important clinical task. Manual examination of colorectal resection slides under a microscope designed for this purpose is time-consuming and requires a high level of expertise. Additionally, access to expert pathologists can be limited, particularly in developing countries or rural settings. Considering the large volume of colonoscopies and CRC screening tests performed each year,^5^ any improvement in the accuracy and/or efficacy of the examination of colorectal resection slides will have a significant impact on public health.

Emerging computer vision and deep learning technologies have led to breakthrough advances in the development of deep learning models, such as convolutional neural networks (CNNs), for histopathology whole-slide image analyses.^6–8^ Such automated models can aid pathologists by identifying slides of interest from a large number of whole slide images (WSIs) for a prioritized review, annotating and augmenting the slides to facilitate the review process, and providing a reliable second opinion if needed. Therefore, the deployment of such models in clinical practice can potentially enhance the accuracy and efficiency of the pathologist’s performance and improve patient outcomes.

There has been ample prior work applying deep learning models for analyzing images related to colorectal cancer. Bychkov et al. used a pretrained network to analyze a single tumor tissue microarray sample from each patient, which generated a probability for the patient’s five-year disease-specific survival.^9^ Xu used a pretrained model to develop a screening tool for pathologists; however, their study only classified a WSI as either cancerous or normal. ^10^ Ho et al. trained a pretrained model to perform strongly supervised glandular segmentations and trained a classical machine learning algorithm to classify the slide as ‘low risk’ or ‘high risk’. ^11^ Choi et al. jointly trained three models on white-light colonoscopic images and used soft ensembling to classify the images into benign, LGD, HGD and adenocarcinoma. ^12^ However, to the best of our knowledge, no other paper has studied the performance of a deep learning model in classifying hematoxylin and eosin (H&E)-stained colorectal resection WSIs into the four classes of benign, LGD, HGD and adenocarcinoma for the purpose of developing a prescreening tool for pathologists.

Dysplasia grading is a complex task, as it has high interobserver variability among pathologists. ^13^ In addition, it is clinically significant, as detecting the existence of an abnormality and grading its extent impact the outcomes of patients who are at high risk of developing colorectal cancer or who have already developed CRC. ^14^ LGD and HGD differ in their architectural features (gland morphology and placement) and cytological features (cell level characteristics). For example, both LGD and HGD display gland crowding but the latter shows back-to-back cribriforming, while the former does not. While both LGD and HGD contain enlarged nuclei, only HGD shows a loss of cell polarity. Current dysplasia grades exist on a sliding scale, and differentiating the intermediate cases at the boundary of LGD and HGD, as well as at the boundary of HGD and adenocarcinoma, can be difficult. ^13^

Considering this complexity and significance, in our study, we develop and evaluate a CNN-based strongly supervised deep learning model for the classification of colorectal surgical resection slides into four classes, adenocarcinoma, HGD, LGD and benign. An automated model that can perform dysplasia grading with high accuracy and with high sensitivity for the high-risk classes of HGD and adenocarcinoma, is a novel addition to the growing number of AI-augmented systems in digital pathology. To demonstrate the model’s generalizability across various institutions with different patient cohorts, devices and data preparation and acquisition protocols, we evaluate our model on a diverse publicly available dataset, in addition to slides from our tertiary medical institution.

## Material and Methods

### Dataset

A total of 889 H&E-stained WSIs were randomly collected from patients who underwent colorectal resections at Dartmouth-Hitchcock Medical Center (DHMC) from 2016 to 2020. In this study, we consider cases with sessile-serrated, tubular, or tubulovillous/villous polyps as LGDs, while slides of normal colonic mucosa or hyperplastic polyps were labeled benign. The polyp types, dysplasia grades, and the presence of adenocarcinoma were extracted according to the associated pathology reports for these slides. Slides were digitized using AT2 scanners (Leica Biosystems, Wetzlar, Germany) at 20x magnification (0.50 μm/pixel). Collected whole-slide images do not overlap with each other, and each slide belongs to a different patient and a separate colonoscopy procedure. This dataset was split into train/validation/test splits, resulting in a training set of 490 slides, a validation set of 165 slides and a test set of 234 slides.

For the purposes of training a strongly supervised model, the regions of interest (ROIs) for the slides in the training and validation sets were annotated. A senior board-certified gastrointestinal (GI) pathologist (A.S.) with over 25 years of experience in gastrointestinal pathology from the Department of Pathology and Laboratory Medicine at DHMC manually annotated the whole-slide images in our training and validation sets. In this annotation process, bounding boxes outlining ROIs for each class were generated using the Automated Slide Analysis Platform (ASAP), a fast viewer and annotation tool for high-resolution histopathology images.^15^

In addition, we collected 606 whole-slide images of CRC from TCGA for external validation. A summary of the distribution of the data across classes and across splits can be found in Table 1.

**Table 1.**
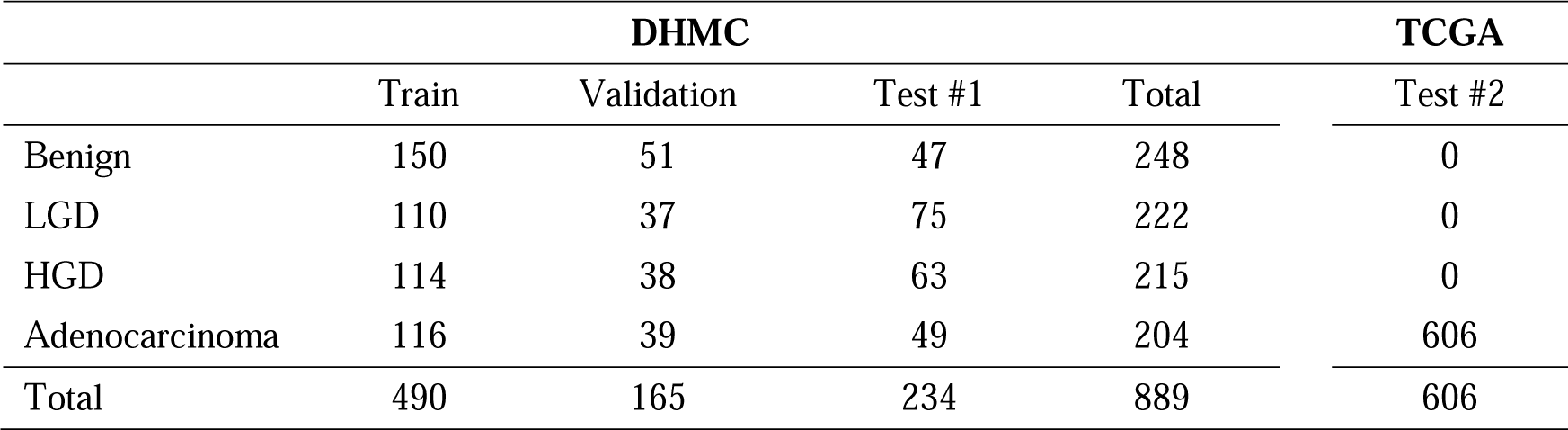
Data distribution of the whole-slide images across all classes and all train/validation/test splits in our datasets.

### Patch extraction and normalization

Based on our preliminary studies^16,17,8,18,19^ and consultations with GI pathology experts, the whole-slide images in our dataset were downsampled to 2.66x magnification (3.75 μm/pixel), as this resolution is sufficient to see clear nuclei structures while allowing the computational units in the CNN model to retrain an effective receptive field. Then, we removed the background from the slides using the tissueloc package^20^ and extracted patches of size 224 × 224 pixels from each WSI. Patches were extracted with no overlap because no significant performance gain was observed in our preliminary experiments when they were extracted with a 1/2 and 1/3 overlap between the patches. The distribution of patches across different classes in our datasets is presented in Table 2. The extracted patches were then Z score normalized by the channelwise mean and standard deviation over all the samples in the training set to account for the staining variations across the slides as well as allowing for stable downstream training and faster convergence.^21^ Figure 1 shows sample patches randomly selected from each class.

**Table 2.** Patch distribution across data splits and classes for the internal dataset and the external test set.

|  | DHMC |  |  |  | TCGA |
| --- | --- | --- | --- | --- | --- |
|  | Train | Validation | Test #1 | Total | Test #2 |
| Benign | 11454 | 3920 | 14401 | 29775 | 0 |
| LGD | 13221 | 1804 | 16516 | 31541 | 0 |
| HGD | 15449 | 5798 | 57502 | 78749 | 0 |
| Adenocarcinoma | 8669 | 2324 | 34070 | 45063 | 464216 |
| Total | 48793 | 13846 | 122489 | 185128 | 464216 |

**Table 3.** Major metrics (%) obtained by our model on the internal test set. The numbers inside indicate (low, high) their 95% confidence intervals.

|  | Accuracy | Sensitivity | Specificity | AUROC |
| --- | --- | --- | --- | --- |
| **Benign** | 96.2 (93.6, 98.3) | 91.7 (83.3, 98.1) | 97.3 (94.8, 99.5) | 97.6 (89.8, 100.0) |
| **LGD** | 98.3 (96.6, 99.6) | 100.0 (100.0, 100.0) | 97.5 (95.0, 99.4) | 99.3 (97.2, 100.0) |
| **HGD** | 93.2 (89.7, 96.2) | 76.3 (65.1, 86.9) | 99.4 (98.2, 100.0) | 97.2 (94.7, 99.4) |
| **Adenocarcinoma** | 94.5 (91.5, 97.0) | 95.9 (89.5, 100.0) | 94.1 (90.5, 97.3) | 98.8 (97.1, 99.9) |
| **Overall** | 95.5 (93.6, 97.4) | 91.0 (87.1, 94.3) | 97.1 (95.9, 98.3) | 98.2 (95.5, 99.6) |

**Figure 1.**
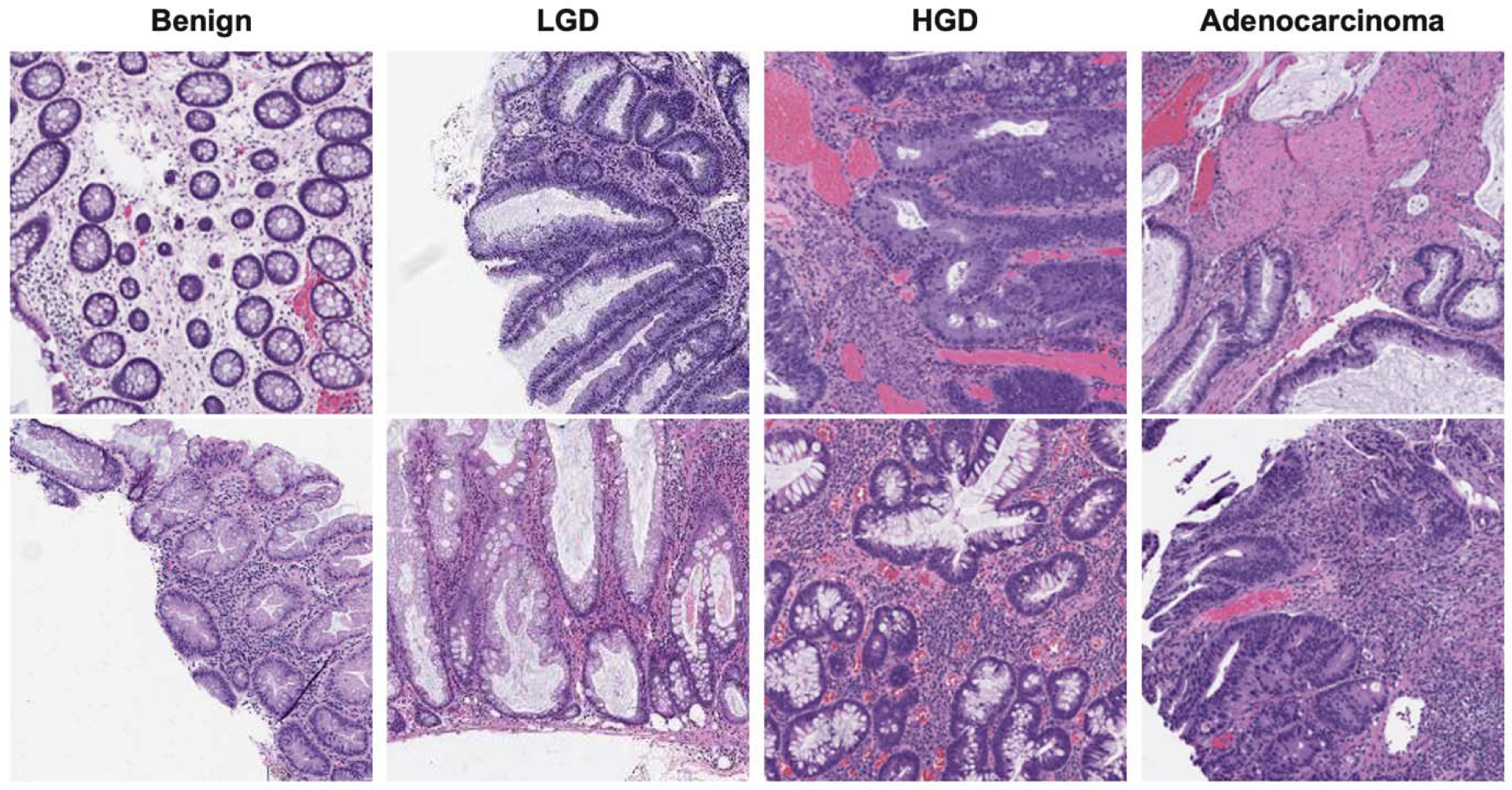
Example extracted patches from each class. From left to right represent benign, LGD, HGD, and adenocarcinoma classes.

### Model development

Our CNN model in this work is a residual neural network with 18 convolutional layers (ResNet-18)^22^. This model was chosen because it demonstrates a high performance over a wide variety of tasks and datasets, including the ImageNet^23^ and COCO^24^ datasets, as well as numerous histopathology datasets.^7,8,25^ A standard ResNet-18 model takes patches of size 224 × 224 pixels as input and outputs probabilities for 1000 classes; therefore, for our purposes, the last classification layer was replaced with output probabilities for the four classes in our dataset (i.e., adenocarcinoma, HGD, LGD and benign). We employed a ResNet-18 model pretrained on the ImageNet dataset, as this led to better validation results due to transfer learning. Furthermore, prior to feeding the patches into the model, we performed a series of data augmentations consisting of random horizontal flipping, random vertical flipping, random rotations and color jittering to improve the generalizability of the model. Random horizontal and vertical flipping is performed with a 50% probability of occurrence. For random rotations, the patches were rotated according to a random sample from choices of 0, 90, 180 and 270 degrees. For color jittering, the brightness, contrast, saturation and hue of each patch are altered by a small amount with a 50% probability of occurrence. The probabilities were chosen to maximize the variety of patches seen during training and were confirmed to be effective against overfitting through cross-validation.

The model was trained by optimizing a multiclass weighted cross-entropy loss function to account for class imbalance. The learning rate was automatically adjusted using a cosine annealing schedule through optimization with an initial learning rate of 0.0001. The Adam optimizer^26^ with an L2 weight decay regularization of 0.0001 was used in our model training. The model was trained for 50 epochs with a batch size of 64 on a Titan Xp graphics processing unit (Nvidia, Santa Clara, CA), which took 2 hours. An illustration of the training pipeline is shown in Figure 2.

**Figure 2.**
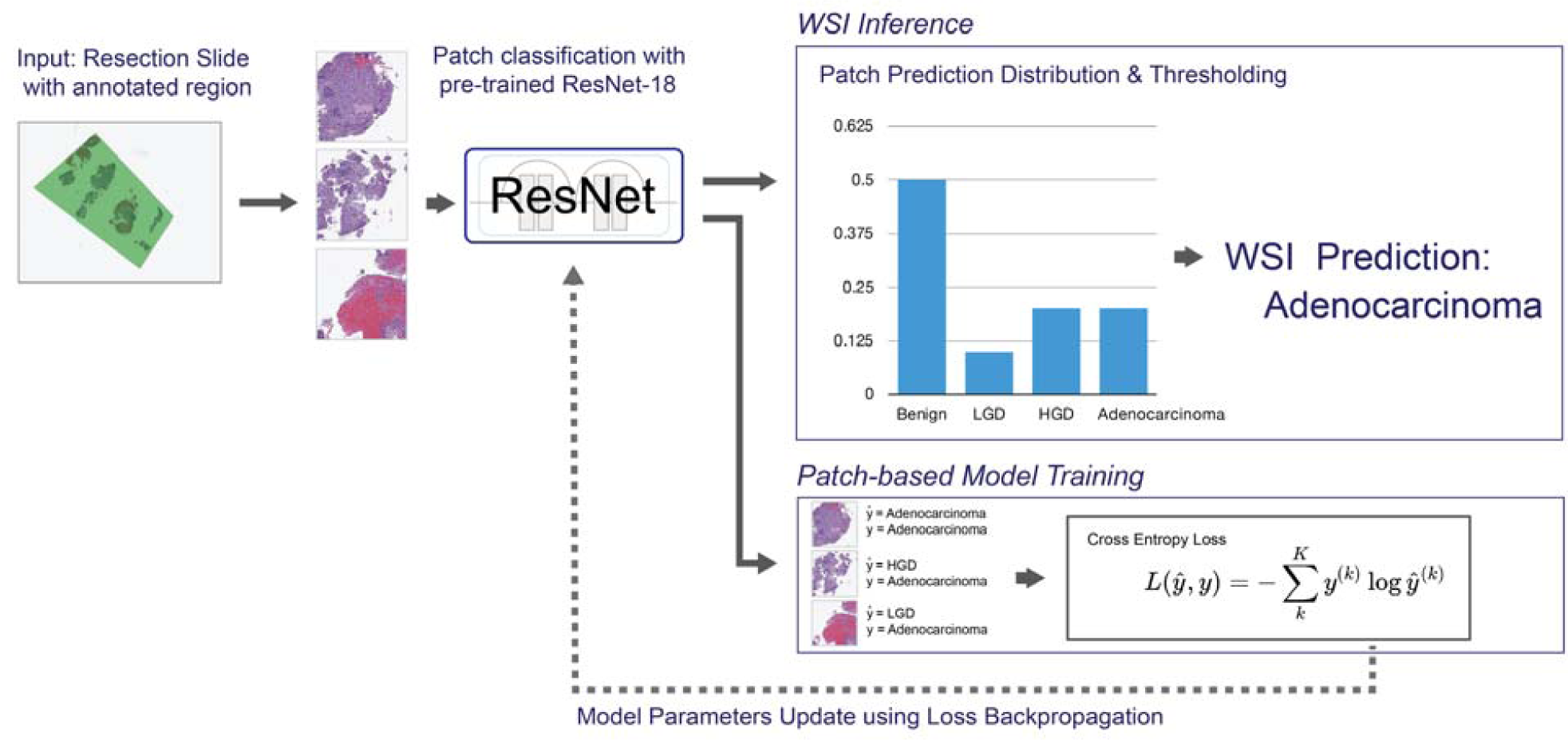
Overview of the training/inference pipeline. The top divergent path is WSI inference; the bottom path is training. Patches are extracted and passed to a ResNet-18 for classification. For training, the class probabilities of a patch calculated by ResNet-18 are used for model optimization through cross-entropy loss backpropagation. During WSI inference, patch prediction distribution was used to deduce the final classification.

### Whole-slide inference and evaluation method

During WSI inference, the patches are extracted from the entire WSI and fed to the trained ResNet-18 to output class predictions for each individual patch. The predictions are aggregated, and the distribution of the predicted patches is passed to a decision tree, which outputs the final prediction for a WSI. The decision tree is constructed as follows: If more than 15% of the patches are predicted as adenocarcinoma, the WSI is classified as adenocarcinoma; otherwise, if more than 10% of the patches are predicted as HGD, the WSI is classified as HGD; otherwise, if more than 5% of the patches are predicted as LGD, the WSI is classified as LGD; otherwise, the WSI is classified as benign. The hierarchy of classes in this decision tree, adenocarcinoma as the most important, benign as least important, allows the model to pick up on important clinical signals. For example, even if only 16% of the patches are cancerous and the rest were benign, the model would still predict that the WSI is adenocarcinoma. The thresholds in this decision tree are fine-tuned using grid search on the validation set, and the final algorithm was reviewed and confirmed by our senior GI pathologist expert. The entire WSI processing and inference for a single WSI takes approximately 1 second in our pipeline.

## Results

Figure 3 shows the confusion matrix, and Figure 4 shows the ROC curves for the model when evaluated on our internal test set. Table 4 reports the accuracy, sensitivity, specificity, and AUROC metrics, as well as their macroaverage values, along with their 95% confidence intervals obtained using bootstrapping^27^ on our internal test set.

**Figure 3.**
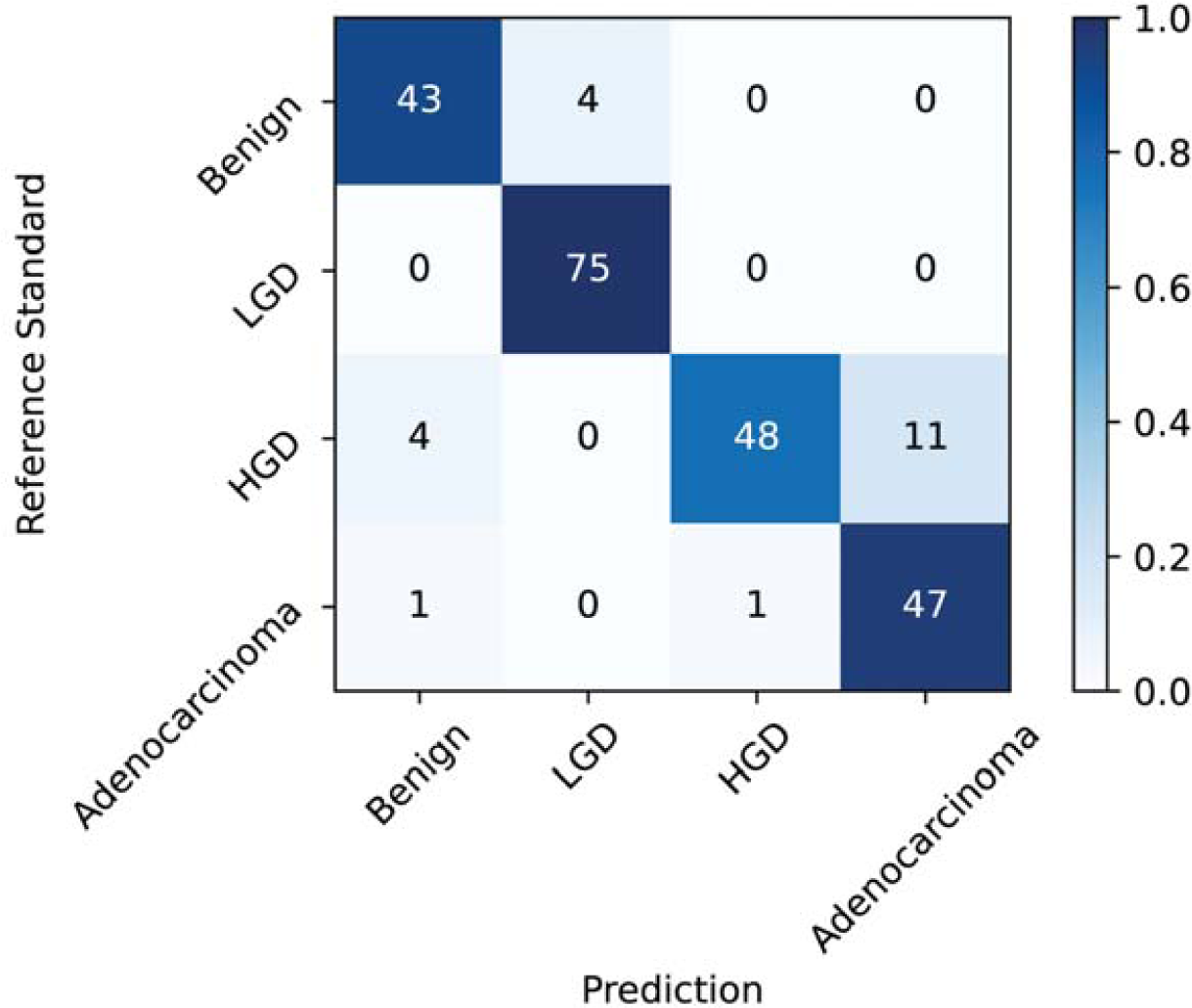
Confusion matrix for the internal test set.

**Figure 4.**
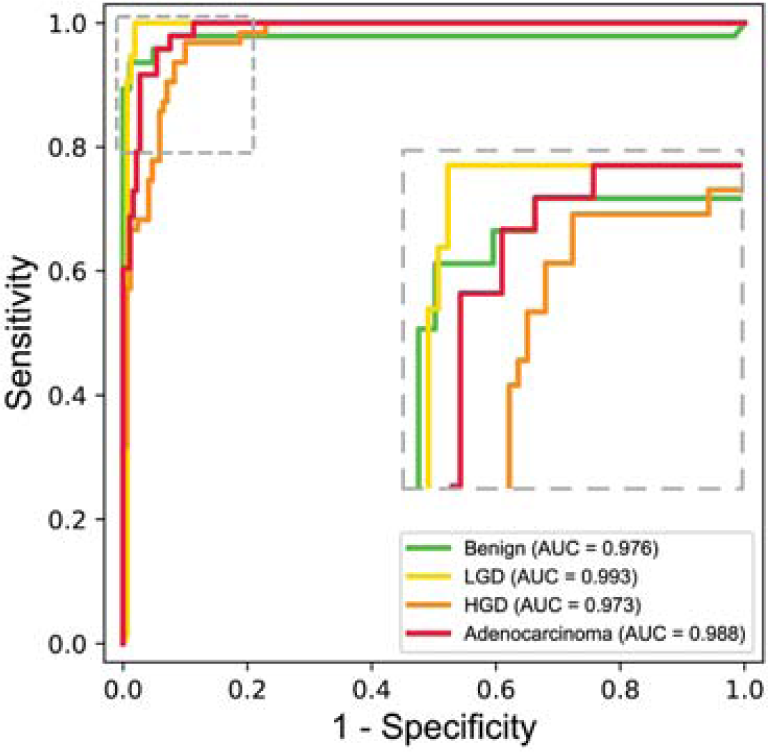
The receiver-operating characteristic (ROC) curves, and the corresponding areas under the ROC curve (AUROC) values, for each of the four classes.

The public TCGA dataset has 606 slides, of which all are adenocarcinoma. On this dataset, our model had an accuracy and sensitivity of 98.5%, with a 95% confidence interval of 97.5% – 99.3%. Since there is only one class (i.e., adenocarcinoma) in this dataset, the accuracy and sensitivity metrics are identical, while the specificity and AUROC could not be computed as meaningful evaluation metrics.

## Discussion

Timely detection of colorectal cancer and dysplasia grading is critical for cancer treatment and prevention; however, it requires a high level of expertise. Deep learning has proven to be useful as a clinical decision support system to assist pathologists in reviewing histopathology slides. The presented, strongly supervised, deep learning model, trained on expert-annotated slides, can assist pathologists in identifying adenocarcinoma on histology slides and quantifying the dysplasia grade.

Our model achieved a promising performance on the adenocarcinoma and HGD cases, with AUROCs of 98.8% (CI: 97.1, 99.9) and 97.2% (CI: 94.7, 99.4), respectively. The sensitivity of our model on adenocarcinoma cases was 95.9% (CI: 89.5, 100.0). This suggests that our model has a low miss rate for cancer cases, which is clinically important. However, we also observed a relatively high level of overcalling of the HGD cases as adenocarcinoma by our model. This effect can be seen in the sensitivity of HGD of 76.3% (CI: 65.1, 86.9), as well as in the confusion matrix in Figure 3, where 11 of the 63 HGD cases were predicted as adenocarcinoma. One adenocarcinoma WSI in our internal test set was predicted to be benign, which can be of concern. However, inspection of the patch-level confidence scores showed that the model correctly identified many adenocarcinoma patches in the slides and considered the case suspicious for cancer but did not have enough evidence in terms of the number of adenocarcinoma patches. Analysis of the patch predictions overlaid on the WSI also shows that the model correctly identifies the cancerous region (Figure 5). Meanwhile, performance on the LGD and benign lesions was extremely high, with AUROCs of 99.3% (CI: 97.2, 100.0) and 97.6% (CI: 89.8, 100.0), respectively. Notably, the model has a sensitivity of 100.0% on LGD.

**Figure 5.**
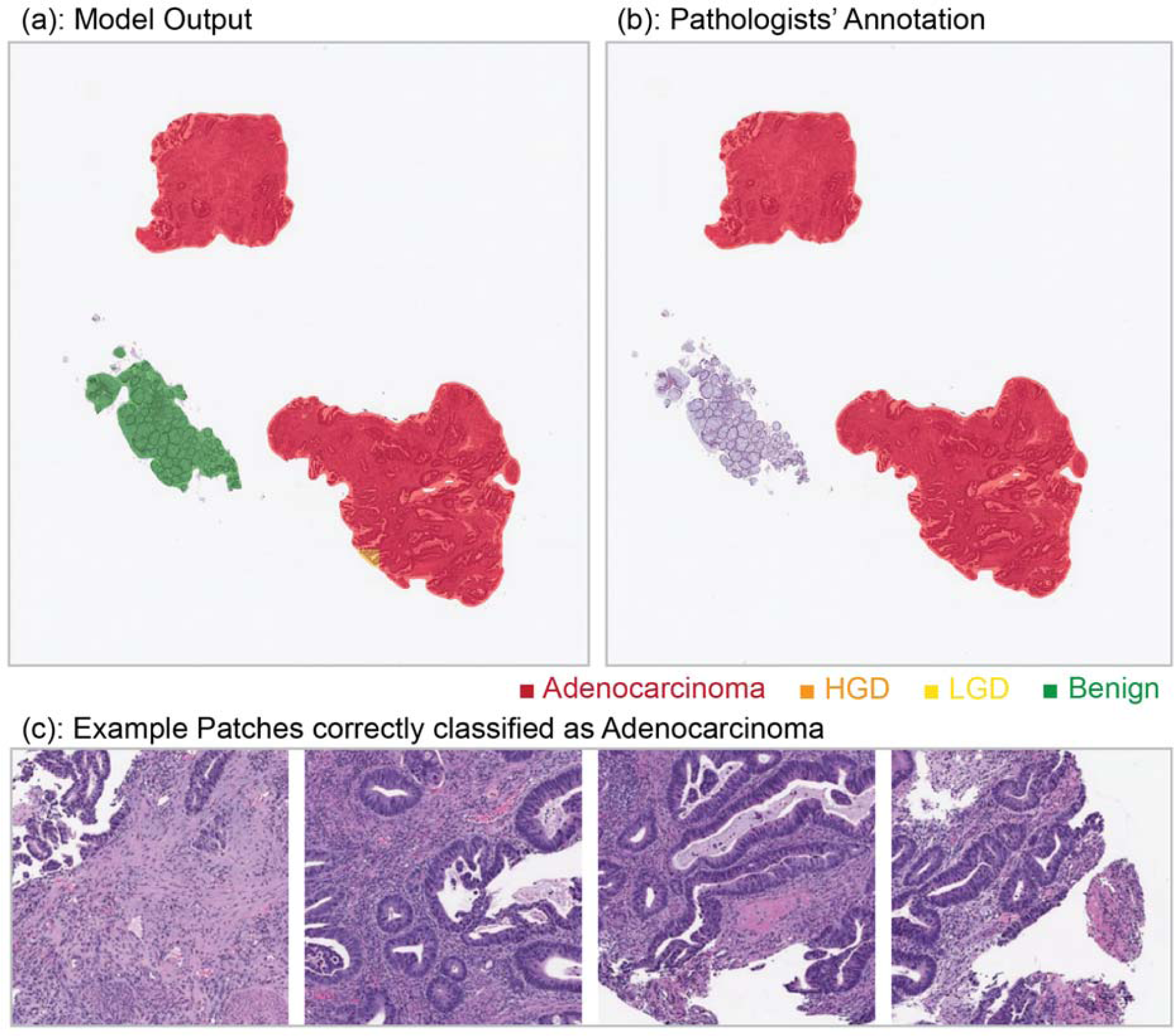
Example adenocarcinoma WSI. (a) Highlighted regions for each class by our model. (b) Annotated ROI by the pathologist. (c) Example patches that are classified as adenocarcinoma by our model.

To perform an error analysis, we presented the model’s results on the internal test set to our senior GI pathology expert (A.S.) at DHMC to identify the source of the discrepancies between the ground truth labels and model predictions. Our GI pathologist concluded that six slides out of the 11 HGD cases that were labeled adenocarcinoma by the model can also be argued to be cancerous, with several suspicious and borderline cancerous regions. Additionally, two out of the remaining five cases had predictions that were on the decision boundary for HGD and adenocarcinoma. Out of the four slides predicted as LGD with a ground truth label of benign, the pathologist also concluded that one slide was in fact LGD. Therefore, out of the 19 misclassifications, seven of them either were suspected correct classifications, while three of the remaining 12 were cases close to the thresholds in the WSI inference decision tree, which could potentially have been solved with extended training and validation sets. On the TCGA set, four out of the seven misclassifications were on the decision boundary of its correct class but fell into a wrong class by a very small margin.

To visually interpret the performance of the model, we overlaid the model predictions on the WSIs and compared them to expert annotations. These visualizations for three slides from adenocarcinoma (Figure 5), HGD (Figure 6) and LGD (Figure 7) classes in the test set are presented below. For each figure, we show the WSI with model prediction overlays, pathologist annotations and example patches from each class.

**Figure 6:**
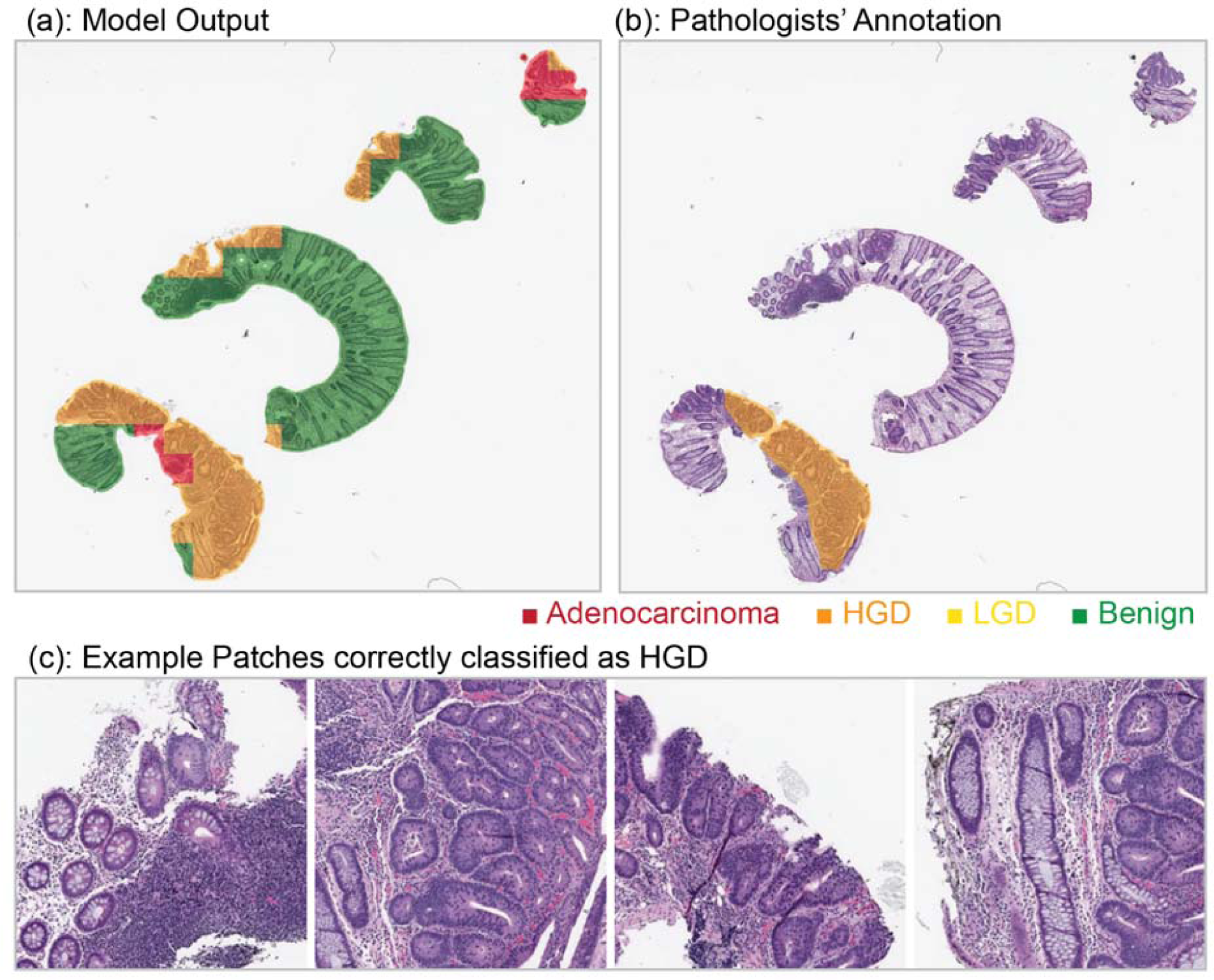
Example HGD WSI. (a) Highlighted regions for each class by our model. (b) Annotated ROI by the pathologist. (c) Example patches that are classified as HGD by our model. Although small regions are suspected to be cancerous by our model, the overall proportion of adenocarcinoma is less than an overall threshold for adenocarcinoma and are treated as outliers.

**Figure 6:**
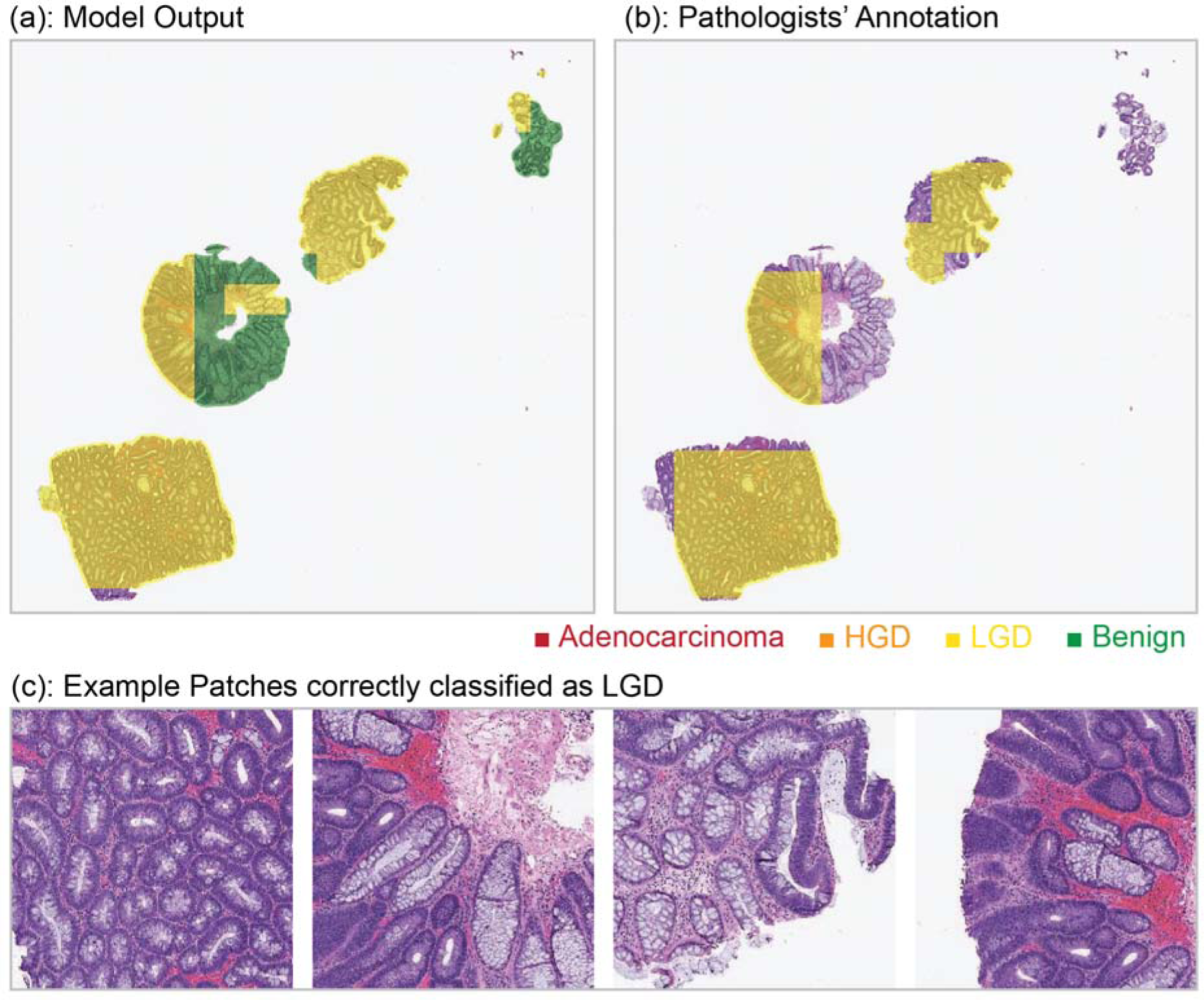
Example LGD WSI. (a) Highlighted regions for each class by our model. (b) Annotated ROI by the pathologist. (c) Example patches that are classified as LGD by our model.

One limitation of this study is the small number of training examples in the dataset, which means that there is not a large variety of cases that are “seen” by the model during the training phase. Deep learning models tend to learn what they see during training. Therefore, a small variety in the histological features present in the training set can lead to a gap between the performance of the model on the training set and the test set. Furthermore, our training data were collected from one medical center. That said, we validated our model’s performance in the detection of adenocarcinoma using the TCGA dataset, which includes slides from different institutions. In future work, we will include a more comprehensive dataset spanning multiple institutions to further train and validate the generalizability of this method.

Acquiring pathologist annotations is resource intensive; therefore, we plan to extend our work by leveraging weakly supervised methods that allow deep learning models to train using the WSI level without expert ROI annotations.^29–32^ Preliminary experiments from our group revealed that the MIL algorithm, a common weakly supervised method trained using our internal training set, works well on the internal test set but does not generalize well to the TCGA dataset. This is most likely due to the small size of our training dataset, as typical MIL methods require over a thousand slides for better generalizations and effectively learning the representative features without ROI-based explicit guidance.^33^ Finally, despite the advances in automated and, AI-powered diagnostic tools, some medical professionals are still hesitant to adopt them in everyday clinical practice, as neural networks lack the transparency and interpretability required by physicians to understand the underlying reasoning of these tools and algorithms. For this reason, our whole-slide inference decision tree is relatively simple and comprehendible by human experts. Additionally, we have provided visualizations highlighting the features and regions that contribute to our model’s classification and WSI inference, so clinicians can gain insight into the reasoning of our model and verify its results. As the next steps, further work must be done to explore new ways to make such tools more transparent and interpretable for use by clinicians.^34^ Finally, our team plans to deploy our developed approach as part of a clinical decision-support system in clinical settings and conduct a follow-up prospective clinical trial with appropriate clinical metrics to evaluate the impact of this work on pathologist performance and patient outcomes.

In summary, in this study, we developed a strongly supervised deep-learning model based on the ResNet-18 architecture to identify colorectal adenocarcinoma and quantify the dysplasia grade on the histopathology slides, which achieved a high performance on an internal test set of 234 whole-slide images of colorectal resection slides. Furthermore, the generalizability of our model for adenocarcinoma detection was demonstrated by evaluating it on a public TCGA dataset consisting of 606 whole-slide images. Based on this strong performance, we conclude that our model has the potential to be used as a clinical decision support system in the domain of digital pathology and can aid pathologists in improving their accuracy and efficiency in reviewing colorectal resection slides.

## Data Availability

TCGA data can be downloaded from the websites: https://portal.gdc.cancer.gov/projects/TCGA-COAD and https://portal.gdc.cancer.gov/projects/TCGA-READ. The DHMC dataset used in this study is not publicly available due to patient privacy constraints. An anonymized version of this dataset can be generated and shared upon reasonable request from the corresponding author.

## AUTHOR CONTRIBUTIONS

Concept and design: A.S. and S.H.; Acquisition, analysis, or interpretation of data: J.K., N.T., A.S., and S.H.; Drafting of the manuscript: J.K. and N.T.; Critical revision of the manuscript for important intellectual content: All authors.; Statistical analysis: J.K. and N.T.; Obtained funding: S.H.; Administrative, technical, and material support: S.H.; Supervision: S.H.

